# NIR-SC-UFES: A portable NIR spectral dataset to skin cancer in vivo

**DOI:** 10.1101/2024.11.27.24317165

**Authors:** Pedro Henrique P. da Cunha, Madson P. Zanoni, Francine D. Santos, Isadora Tavares Nascimento, Isabella Rezende, Tania R. P. Canuto, Luciana de Paula Vieira, Renan Rossoni, Maria C. S. Santos, Wanderson Romão, Patricia H. L. Frasson, Renato A. Krohling, Paulo R. Filgueiras

**Author notes:** **Corresponding author’s email address and Twitter handle**.

## Abstract

In recent years, significant progress has been made in computer-aided diagnostics (CAD) for skin lesions, primarily using images and metadata. However, these methods have limitations, particularly in revealing the molecular structure of lesions. NIR spectroscopy offers additional data, capturing information not visible to the naked eye, which can improve automated CAD for skin lesions. Skin cancer remains a major cause of death, making early diagnosis crucial. A key challenge in applying machine and deep learning (MDL) techniques to spectroscopy is the lack of publicly available datasets. Previously, no public dataset of portable NIR spectral data for skin lesions existed. In collaboration with the Programa de Assistência Dermatológica (PAD) at UFES, we developed a new dataset, NIR-SC-UFES, for skin cancer diagnosis. This dataset includes portable NIR spectral data for six lesion skin, with 714 spectra captured between 900 and 1700 nm. The dataset is available at data.mendeley.com/datasets/j9773cyr3k/1.

**Graphical abstract:** 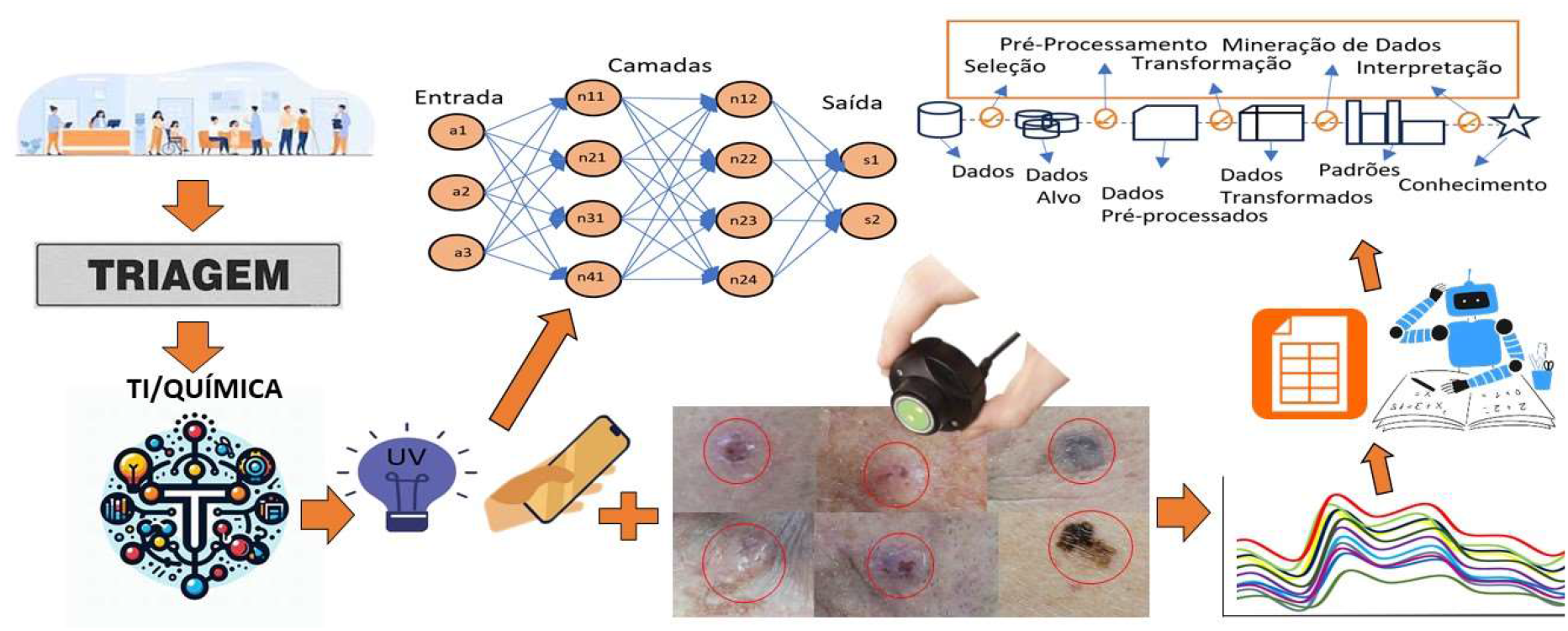

**Specifications Table:** 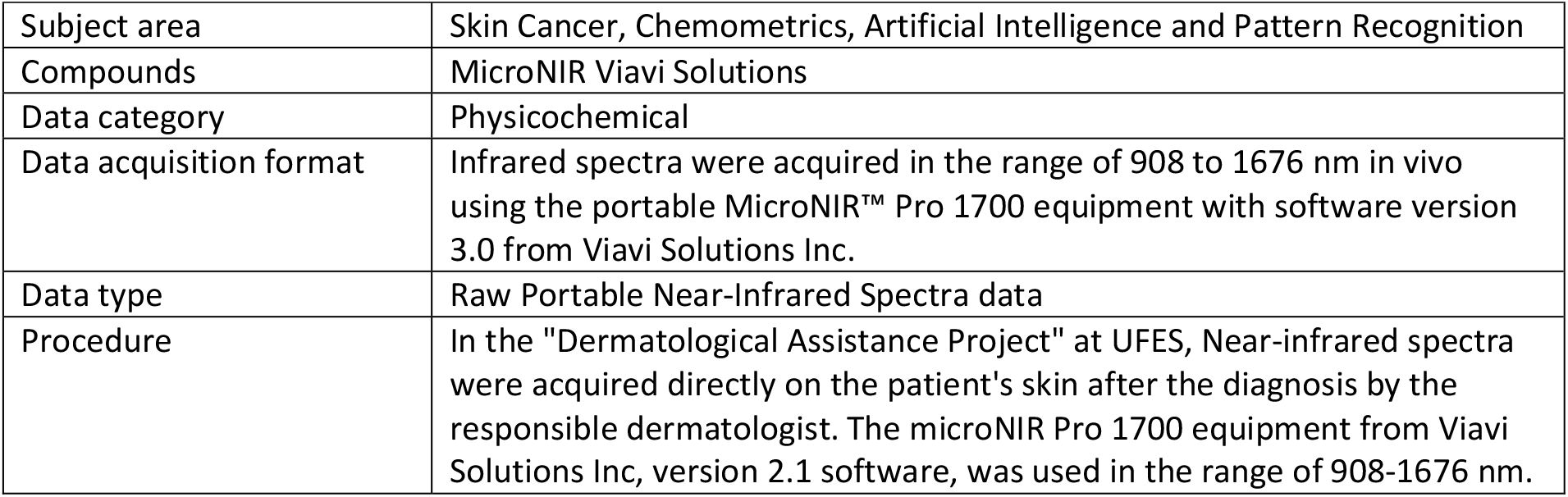

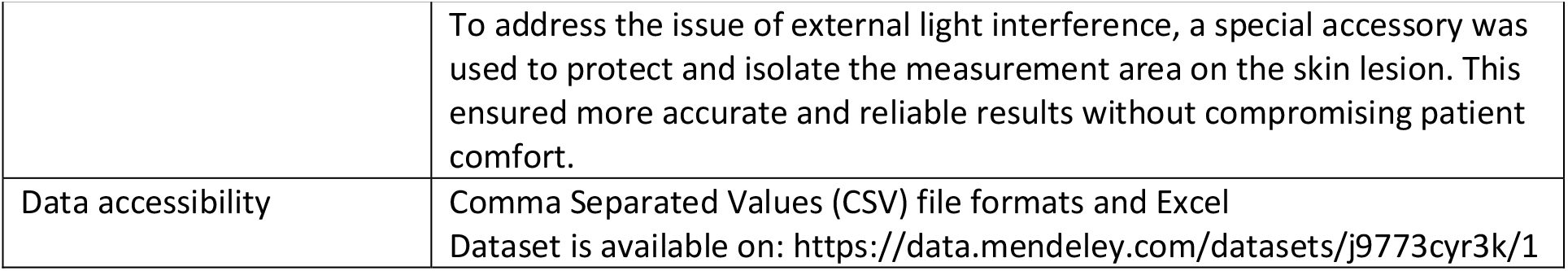

## 1. Rationale

Near Infrared (NIR) spectroscopy is a technique that detects molecular vibrations and provides information about the bonds between atoms. Through NIR spectroscopy, it is possible to understand the chemical bonds present in a sample. This technique, combined with chemometric methods, has been applied in various areas, including plastic waste treatment, food fraud detection, and assessment of skin health [1–3].

The infrared technique offers significant advantages compared to other analysis techniques. It allows for quick, non-destructive analysis and environmentally sustainable. Furthermore, it does not require prior sample preparation [2]. With the advancement of portable versions, although they may have lower sensitivity compared to traditional versions, the technique becomes more versatile and expands its potential for field applications [3,4]. Additionally, microNIR has been applied in the field of human health [5], including dermatology [6].

Skin cancer accounts for approximately one-third of all cancers in Brazil, according to the Brazilian Society of Dermatology. There are six main types of skin lesions: Actinic Keratosis (ACK), Basal Cell Carcinoma (BCC), Malignant Melanoma (MEL), Melanocytic Nevus (NEV), Squamous Cell Carcinoma (SCC), and Seborrheic Keratosis (SEK). Among these, three are benign (ACK, NEV, and SEK) and three are malignant (BCC, MEL, and SCC). These lesions are show in Figure 1. In the case of malignant cancers, immediate removal is recommended due to the high likelihood of progression and metastasis development.

**Figure 1.**
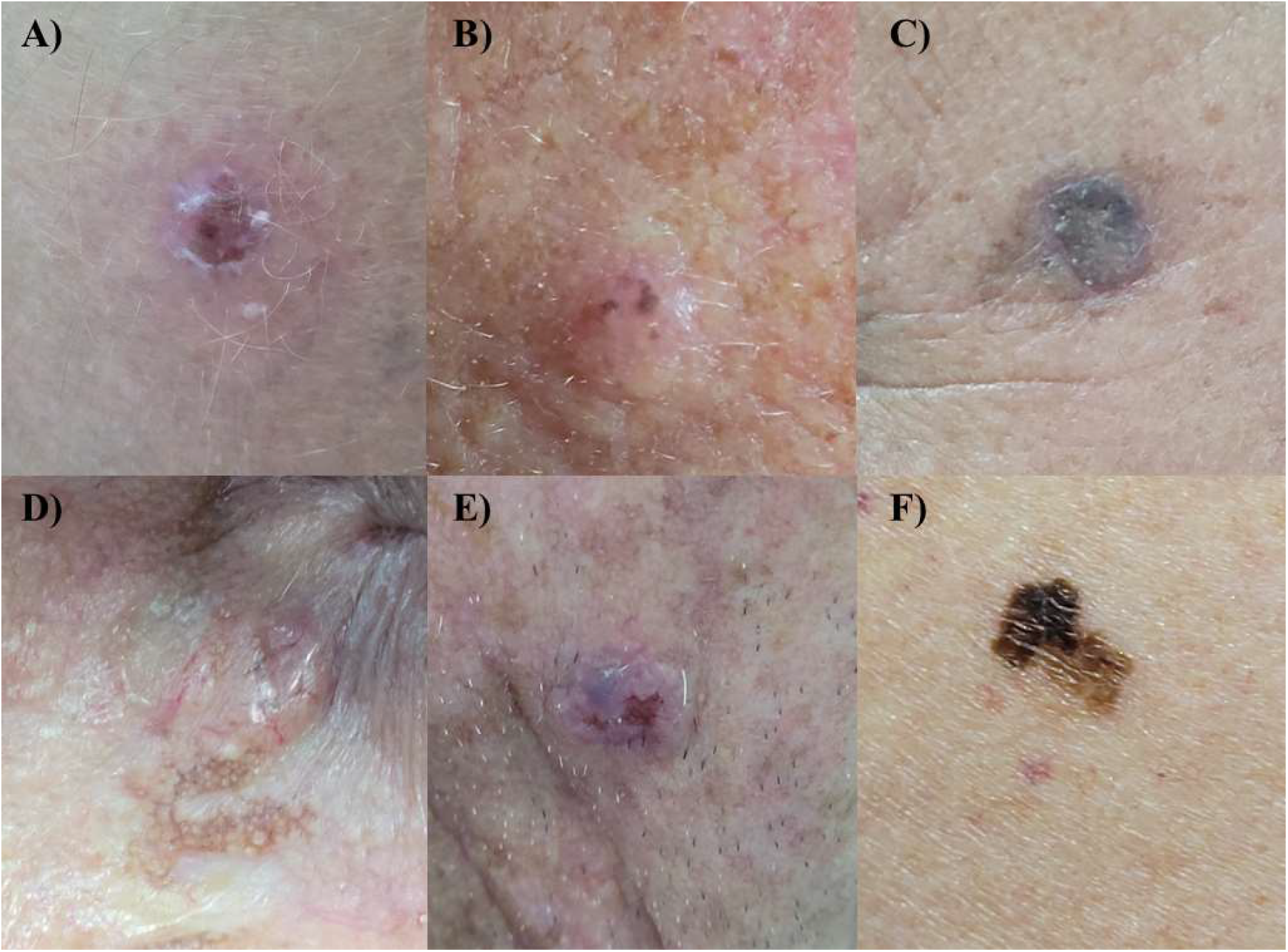
Skin lession. A) Actinic Kerotosis, B) Melanocytic Nevus, C) Seborrheic Keratosis, D) Basal Cell Carcinoma, E) Squamous Cell Carcinoma and F) Malignant Melanoma,

This database contains NIR spectra of 714 dermatological lesions from 331 different patients collected in the PAD over a period of two years. The lesions were classified into six main categories, with the quantities presented in Table 1. It is important to note that there is smaller number of melanoma samples compared to the other lesions.

**Table 1.**
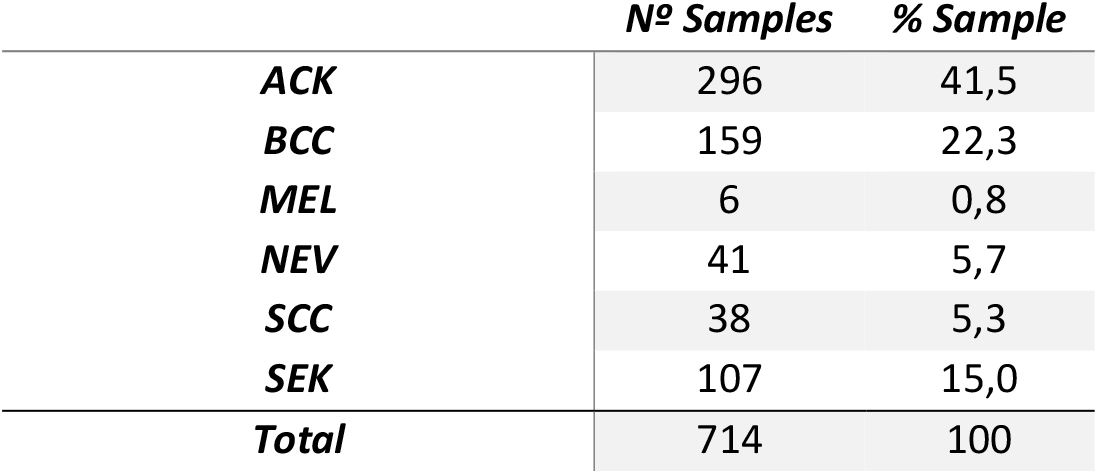
Number of skin lesion samples.

## 2. Procedure

The PAD is an initiative of UFES with the primary aiming to diagnose and treat skin cancer in communities in the interior of Espírito Santo. The program carries out awareness campaigns about skin health among the local population and intervenes in the course of the disease through prevention, diagnosis, and treatment of possible cases of skin cancer. Indeed, the work carried out by PAD is of utmost importance for the communities in the interior, especially for individuals of European descent. Collaboration with the Laboratory of Computing and Nature-Inspired Engineering (LABCIN-UFES) and the Chemometrics Laboratory (Q-UFES), enabled the collection of clinical and spectral data from patients during their consultations.

### 2.1 Equipment description

The NIR spectra were obtained in triplicate in the wavelength range of 908 to 1676 nm. A total of 100 scans were performed with a resolution of 6.20 nm, resulting in a total measurement time of 8 seconds for each sample. The microNIR equipment includes the VIAVI MicroNIR Pro software, as shown in Figure 2, which enables data acquisition, calibration, method development, user management, and real-time predictions.

**Figure 2.**
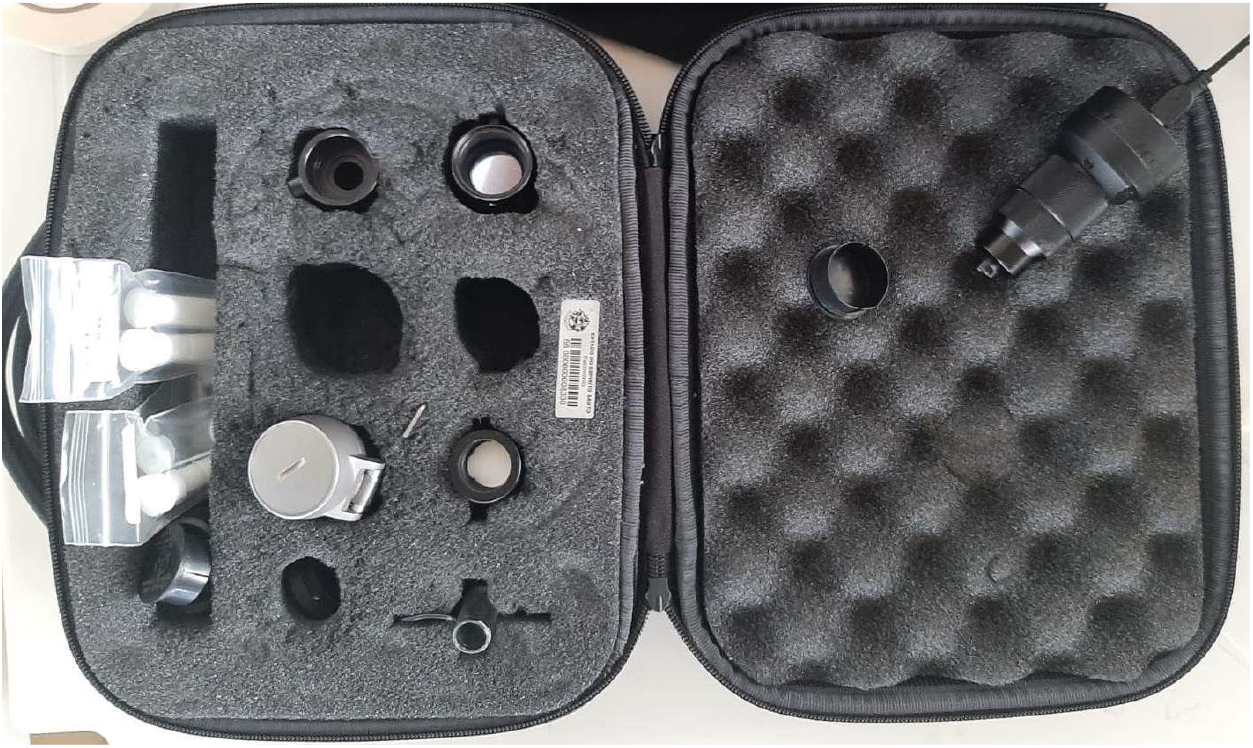
MicroNIR kit and its accessories.

In addition, a conical-shaped accessory with an internal gold sheet was used to assist in collection of spectra. This accessory was attached to the infrared emitter equipment to minimize the loss of light information and prevent undesired noise.

### 2.2 Data Collecion

At the healthcare medical, students perform an initial analysis of patients lesions and refer them to experienced dermatologists for confirmation of the diagnosis. The students are present to learn and apply their knowledge in practice. After the analysis conducted by the dermatology team, patients are referred according to the type of lesion they present. Before any treatment or surgery, patients are referred to the Information Technology (IT) and Chemistry team. This step is crucial for collecting information through images, as well as conducting analyses using the microNIR technique. These analyses are performed on the lesions without any interference from treatment, ensuring more accurate results.

Before collecting the spectra, the equipment performs a scan of the dark environment and the reference to calibrate the device. This is done to minimize the effects of ambient light and reduce undesired noise. After calibration, the patient’s number provided by the National Health Service (Sistema Único de Saúde in Portuguese) is recorded for sample identification. It is important to note that this number is only used for biopsy confirmation and is not present in the datset.

A support is attached to microNIR device to assist in analyses, ensuring that the quartz window of the equipment does not come into direct contact with the injured skin, as shown in Figure 3. The collection is performed on the lesions of interest, including benign lesions (ACK, SEK, and NEV) and malignant lesions (BCC, SCC, and MEL). After analyzing the samples from all the lesions, patients are referred according to the type of lesion.

**Figure 3.**
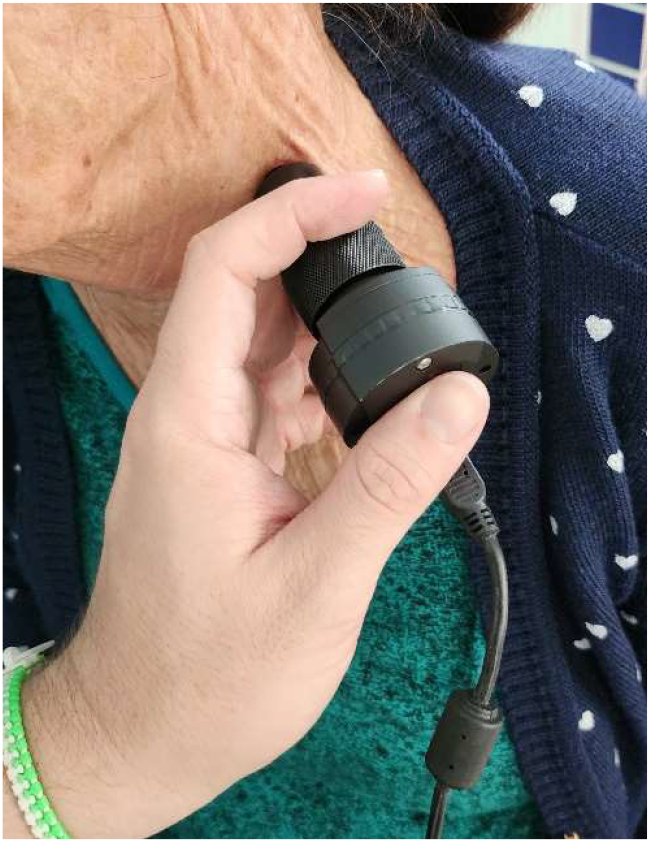
MicroNIR used to collect the dataset NIR-SC-UFES

Benign lesions were treated with specific ointments according to their type. In the case of malignant lesions (BCC, SCC, MEL, and others), the plastic surgery team performed surgical procedures to remove them. The removed lesions were sent for biopsy and further official description in the medical report. The tip of the accessory is cleaned for each analysis on different patients to ensure equipment and patient hygiene. Lesions that exhibited pain sensitivity were not analyzed.

## 3. Data, value and validation

As presented in Table 1, the samples exhibit an imbalance in classes, with a lower percentage of MEL compared to the others and a higher amount of ACK. However, this disparity reflects the naturally occurring distribution in the population, where MEL is rare and ACK is common. One possible strategy to resolve this imbalance is the use of artificially created samples, as demonstrated in the work of Loss F. P. et al., in 2024, which employed the Synthetic Minority Over-sampling Technique (SMOTE) to enhance classification models.

### Ethics statements

Ethics Statement The dataset was collected along with the Dermatological and Surgical Assistance Program (PAD) of the Federal University of Espírito Santo. The program is managed by the Department of Specialized Medicine and was approved by the university ethics committee (nr. 50 0 0 02/478) and the Brazilian government through Plataforma Brasil (nr. 4.007.097), the Brazilian agency responsible for research involving human beings. In addition, all data is collected under patient consent and the patients privacy is completely preserved.

## Data Availability

All dataset is available on: https://data.mendeley.com/datasets/j9773cyr3k/1

## Acknowledgments

This study was financed in part by the Coordenação de Aperfeiçoamento de Pessoal de Nível Superior – Brasil (CAPES) – Finance Code 001; the Conselho Nacional de Desenvolvimento Científico e Tecnólogico (CNPq) – grant n. 309729/2018-1 – and the Fundação de Amparo a Pesquisa e Inovação do Espírito Santo (FAPES) – grant n. 575/2018. We also thank the support of the Secretary of Health of the Espírito Santo state (SESA), the municipal administration of the 11 cities in which PAD takes place, and the Lutheran church of the Espírito Santo state. Lastly, we acknowledge the work of Prof. Carlos Cley and Prof. Luiz F. S. de Barros who founded the PAD in 1987.

## Notes

### Competing Interest Statement

This study was financed in part by the Coordenacao de Aperfeicoamento de Pessoal de Nivel Superior Brasil (CAPES) Finance Code 001; the Conselho Nacional de Desenvolvimento Cientifico e Tecnologico (CNPq) grant n. 309729/2018-1 and the Fundacao de Amparo a Pesquisa e Inovacao do Espirito Santo (FAPES) grant n. 575/2018. We also thank the support of the Secretary of Health of the Espirito Santo state (SESA), the municipal administration of the 11 cities in which PAD takes place, and the Lutheran church of the Espirito Santo state. Lastly, we acknowledge the work of Prof. Carlos Cley and Prof. Luiz F. S. de Barros who founded the PAD in 1987.

### Funding Statement

This study was financed in part by the Coordenacao de Aperfeicoamento de Pessoal de Nivel Superior - Brasil (CAPES) - Finance Code 001; the Conselho Nacional de Desenvolvimento Cientifico e Tecnologico (CNPq) - grant n. 309729/2018-1 - and the Fundacao de Amparo a Pesquisa e Inovacao do Espirito Santo (FAPES) - grant n. 575/2018. We also thank the support of the Secretary of Health of the Espirito Santo state (SESA), the municipal administration of the 11 cities in which PAD takes place, and the Lutheran church of the Espirito Santo state. Lastly, we acknowledge the work of Prof. Carlos Cley and Prof. Luiz F. S. de Barros who founded the PAD in 1987.

### Author Declarations

The dataset was collected along with the Dermatological and Surgical Assistance Program (PAD) of the Federal University of Espirito Santo. The program is managed by the Department of Specialized Medicine and was approved by the university ethics committee (nr. 500002/478) and the Brazilian government through Plataforma Brasil (nr. 4.007.097), the Brazilian agency responsible for research involving human beings. In addition, all data is collected under patient consent, and the patients' privacy is completely preserved.

## References

[1] F.P. Loss, P.H. da Cunha, M.B. Rocha, M.P. Zanoni, L.M. de Lima, I.T. Nascimento, I. Rezende, T.R.P. Canuto, L. de P. Vieira, R. Rossoni, M.C.S. Santos, P.L. Frasson, W. Romão, P.R. Filgueiras, R.A. Krohling, Skin cancer diagnosis using NIR spectroscopy data of skin lesions in vivo using machine learning algorithms, (2024). 10.48550/arXiv.2401.01200.

[2] F.D. Santos, L.P. Santos, P.H.P. Cunha, F.T. Borghi, W. Romão, E.V.R. de Castro, E.C. de Oliveira, P.R. Filgueiras, Discrimination of oils and fuels using a portable NIR spectrometer, Fuel 283 (2021) 118854. 10.1016/j.fuel.2020.118854.

[3] G.S. Folli, L.P. Santos, F.D. Santos, P.H.P. Cunha, I.F. Schaffel, F.T. Borghi, I.H.A.S. Barros, A.A. Pires, A.V.F.N. Ribeiro, W. Romão, P.R. Filgueiras, Food analysis by portable NIR spectrometer, Food Chem. Adv. 1 (2022) 100074. 10.1016/j.focha.2022.100074.

[4] O. Escuredo, M.S. Rodríguez-Flores, L. Meno, M.C. Seijo, Prediction of Physicochemical Properties in Honeys with Portable Near-Infrared (microNIR) Spectroscopy Combined with Multivariate Data Processing, Foods 10 (2021) 317. 10.3390/foods10020317.

[5] R. Risoluti, G. Gullifa, F. Buiarelli, S. Materazzi, Real time detection of amphetamine in oral fluids by MicroNIR/Chemometrics, Talanta 208 (2020) 120456. 10.1016/j.talanta.2019.120456.

[6] L.M. McIntosh, M. Jackson, H.H. Mantsch, J.R. Mansfield, A.N. Crowson, J.W.P. Toole, Near-infrared spectroscopy for dermatological applications, Vib. Spectrosc. 28 (2002) 53–58. 10.1016/S0924-2031(01)00165-5.

